# Preprint server use in kidney disease research: a rapid review

**DOI:** 10.1101/2020.03.16.20036590

**Authors:** Caitlyn Vlasschaert, Cameron Giles, Swapnil Hiremath, Matthew B. Lanktree

## Abstract

**Purpose of review:** Preprint servers including arXiv and bioRxiv have disrupted the scientific communication landscape by providing rapid access to pre--peer reviewed research. MedRxiv is a recently launched free online repository for preprints in the health sciences. We sought to summarize potential benefits and risks to preprint server use, from both the researcher and end--user perspective, and evaluate the uptake of preprint servers in the nephrology community.

**Sources of Information:** We performed a rapid review of articles describing preprint servers and their use. We approached the 20 highest impact nephrology journals regarding their policy towards the use of preprint servers. We evaluated the average time from study completion to publication of impactful articles in nephrology. Finally, we evaluated the number of nephrology articles submitted to preprint servers.

**Findings:** To date over 600 kidney--related articles have been uploaded to bioRxiv and medRxiv. The average time from study completion to publication was over 10 months. 16 of the top 20 nephrology journals currently accept research submitted to a preprint server. Transparency and collaboration, visibility and recognition, and rapid dissemination of results were identified as benefits of preprint servers. Concerns exist regarding the potential risk of non--peer reviewed medical research being publicly available.

**Limitations:** Preprint servers remain a recent phenomenon in health sciences and their long-- term impact on the medical literature remains to be seen.

**Implications:** The quantity of research submitted to preprint servers is likely to continue to grow. The model for dissemination of research results will need to adapt to incorporate preprint servers.

## Introduction

Preprint servers are a disruptive force in scientific communication, bound to alter the models by which medical knowledge is disseminated. Preprints are draft manuscripts presenting original research that have not yet undergone peer review. Since the 1950s, paper preprints have been shared amongst medical colleagues^1^, and even circulated within National Institutes of Health (NIH)--organized networks of like--minded researchers^2^, with a goal of bettering a manuscript prior to submission for formal peer review and evaluation of its suitability for publication by journal editors. Preprint servers present an option for public posting of pre--peer reviewed manuscripts with the goal of establishing a public record of novel ideas, improving the quality of the manuscript before journal submission, and improving the rapidity of research dissemination. Researchers utilizing an electronic preprint server typically upload their work onto a preprint server while simultaneously submitting it to a traditional journal for peer review.

The first widely used electronic preprint server, called arXiv (pronounced “archive”), was developed by the physics and mathematics community in 1991. In the last five years, preprint servers have experienced rapid uptake by the life sciences research community^3^. BioRxiv is a free online preprint server in the life sciences founded in 2013, which hosts over 75,000 preprints as of March 2020. Access to the server is growing exponentially: there were over 2.2 million preprint downloads for October and November 2018, which is more than the cumulative total of downloads during its first 2.5 years of operation^4^. More than 170 journals have partnered with bioRxiv to create a streamlined submission process called “B2J”, where manuscripts can be directly transferred to the peer--reviewed journals at the time of preprint upload^5,6^.

Adoption of preprint servers has lagged in medicine compared to other research fields, at least in part because many major medical journals have, until recently, refused to publish preprinted papers^7,8^. The preprint culture in medicine is evolving: many medical journals have reversed their historic policies and now accept preprints^9,10,11^. MedRxiv is a medicine--specific preprint repository launched in June 2019. Its stated goals are to “improve the openness and accessibility of scientific findings, enhance collaboration among researchers, document the provenance of ideas, and inform ongoing and planned research through more timely reporting of completed research”^12^. However, concerns about possible dangers of early public access to non--peer reviewed medical research remain^13,14^. Preprints are beginning to provide a means for timely discussion of medical research, complementary to other non--peer reviewed modalities such as conference presentations and educational blog posts. With one of the most interactive and engaged online medical communities^15,16^, the field of nephrology could have a large influence on the uptake of preprints in medicine. This rapid review will evaluate potential risks and benefits associated with preprint server use from the author, reader, journal and granting agency perspective and evaluate the current state of preprint server utilization in nephrology.

## Methods

We performed a modified rapid review to evaluate the role of preprint servers in kidney research. We searched the term “preprint” on PubMed on December 20^th^, 2019 and imported all results into Covidence, a software platform that streamlines article screening and data extraction for systematic reviews^17^. Two independent reviewers (CV & CG) screened all texts for relevance to this article.

To evaluate the uptake of preprint server use in the nephrology field, we searched the titles and abstracts of bioRxiv and medRxiv preprint uploads on January 20, 2020 using the following keywords: “kidney” or “kidneys” or “renal” or “nephrology” or “nephron” or “nephropathy” or “nephrotic” or “nephritic” or “nephrolithiasis” or “ESRD” or “chronic kidney disease” or “CKD” or “dialysis” or “electrolyte” or “hypertension” or “urinalysis” or “glomerulus” or “hypokalemia” or “hyperkalemia” or “hypernatremia” or “hyponatremia” or “hypocalcemia” or “hypercalcemia” or “hepatorenal” or “cardiorenal” or “glomerulonephritis” or “GFR”. This returned 1117 results for bioRxiv and 277 results for medRxiv. The article titles and abstracts were screened for relevance to the field of nephrology by two independent reviewers (CV & CS). In total, 628 bioRxiv preprints (504 basic science and 124 clinical research) and 39 medRxiv preprints were deemed relevant and included in the analysis in Figure 1. Next, we calculated the time from study completion to publication for the most impactful clinical trials in nephrology, as determined by a Twitter poll (https://twitter.com/Renaltubules/status/1233565802768543746). Twenty--seven studies were included in the analysis and data regarding study completion and publication dates was obtained from the seminal publication and trial registry websites (www.clinicaltrials.org). Finally, we contacted the managing editors for the 20 highest impact journals in nephrology according to the Scimago Journal & Country Rank list in order to inquire about their policy on preprinting.

**Figure 1.**
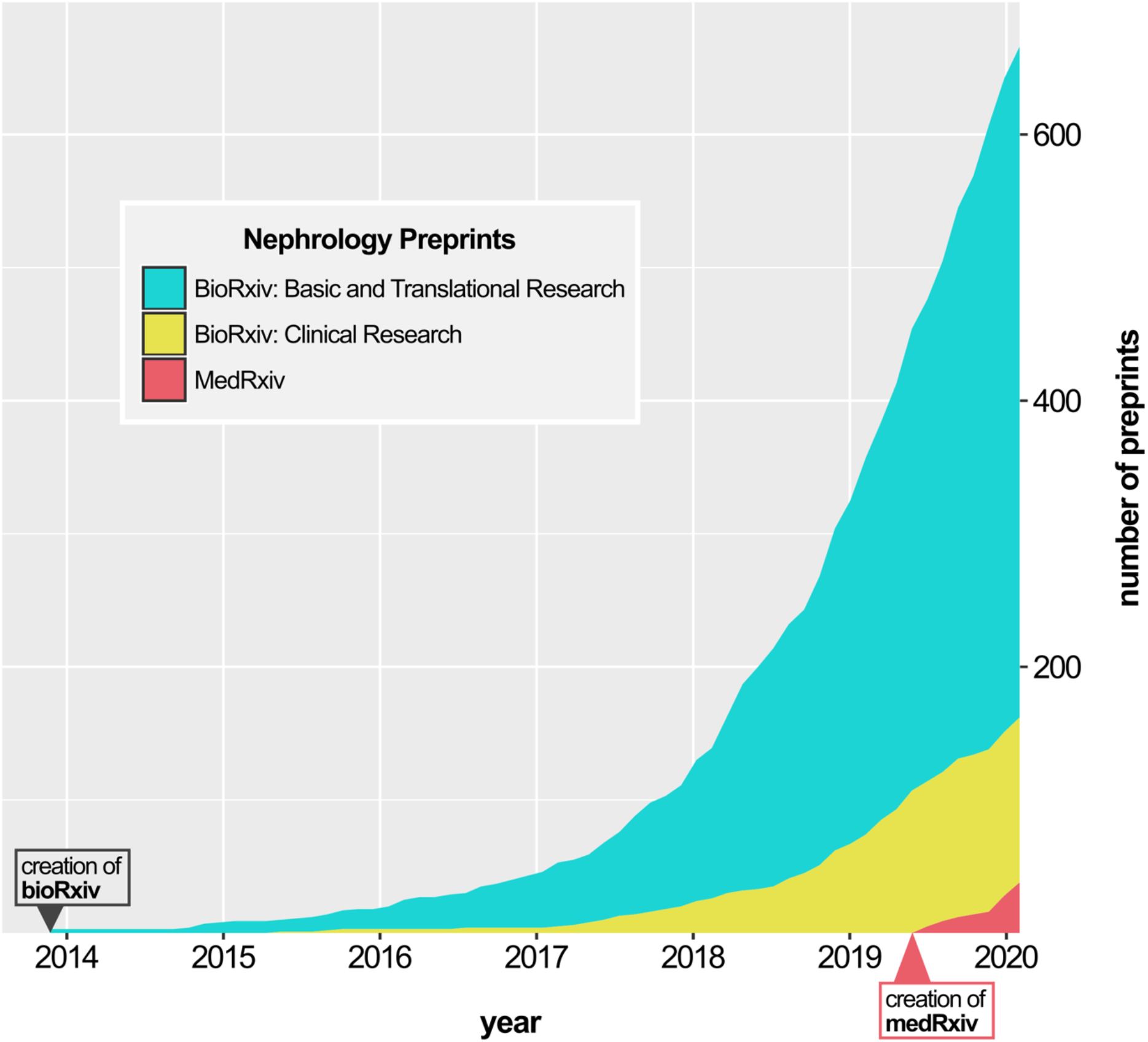
Rapid growth in nephrology preprint papers in bioRxiv and medRxiv since 2013.

**Figure 2.**
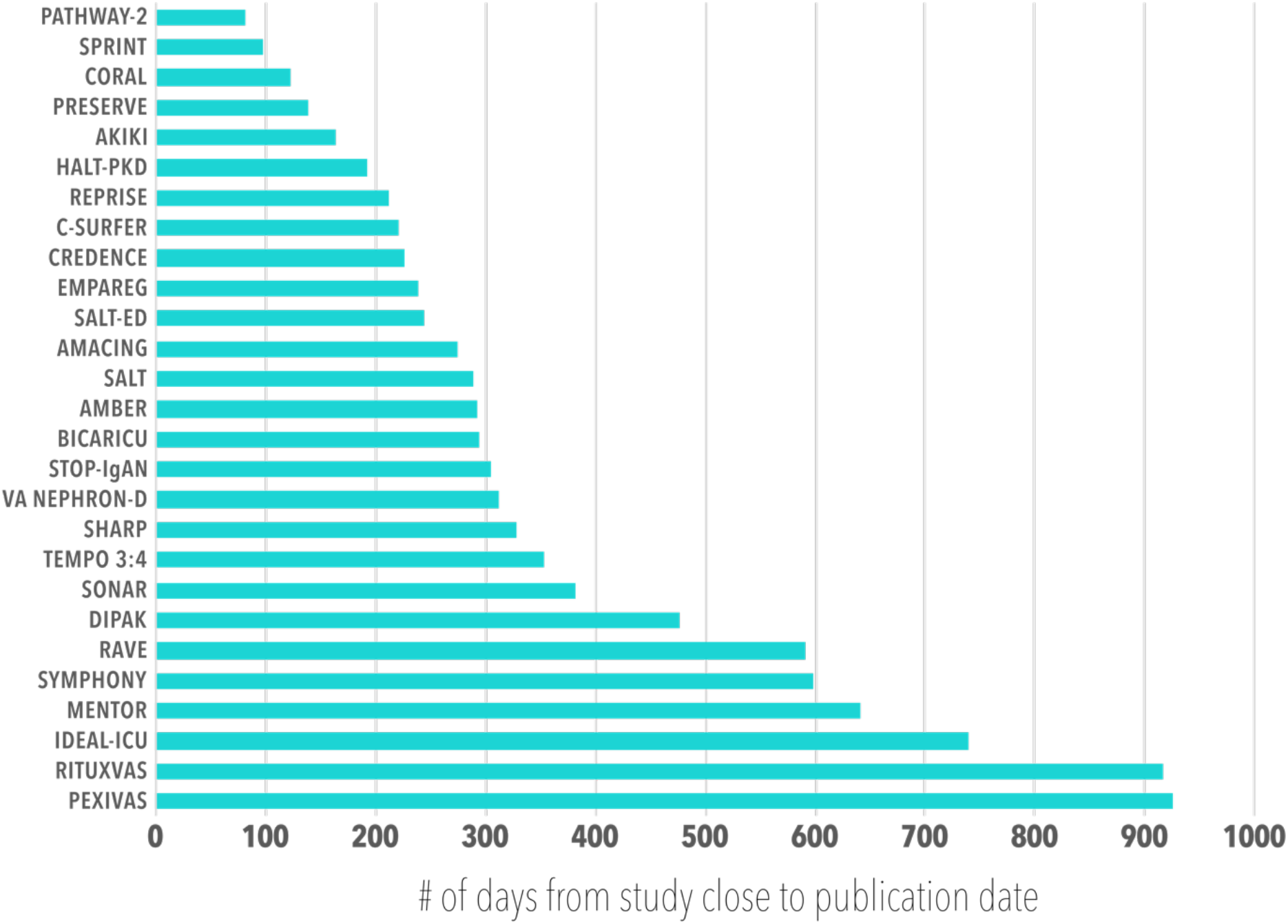
Elapsed time between study close to publication date for impactful nephrology clinical trials (average: 10.44 months).

## Results

### Preprints in nephrology: current uptake

The field of nephrology has a large online presence with prominent post--publication discussions in the form of journal clubs (i.e., the Twitter--based NephJC^18^) and blog posts (e.g., on the Renal Fellow Network^19^ and the American Journal of Kidney Disease (AJKD) blog^20^). Preprints are making an exponential appearance in the workflow of nephrology research: to date, over 600 kidney--related manuscripts have been uploaded to preprint servers (Figure 1). This includes 124 clinical papers in biorXiv, 504 basic science papers in bioRxiv, and 39 clinical papers in medRxiv as of January 20, 2020.

To evaluate the opportunity for faster research dissemination in nephrology, we collected the length of time between study completion and publication of the most impactful publications in nephrology. These trials took 10.4 months on average from completion to publication, ranging from 2 months and 11 days to 2 years, 6 months and 16 days (Figure 3).

**Figure 3.**
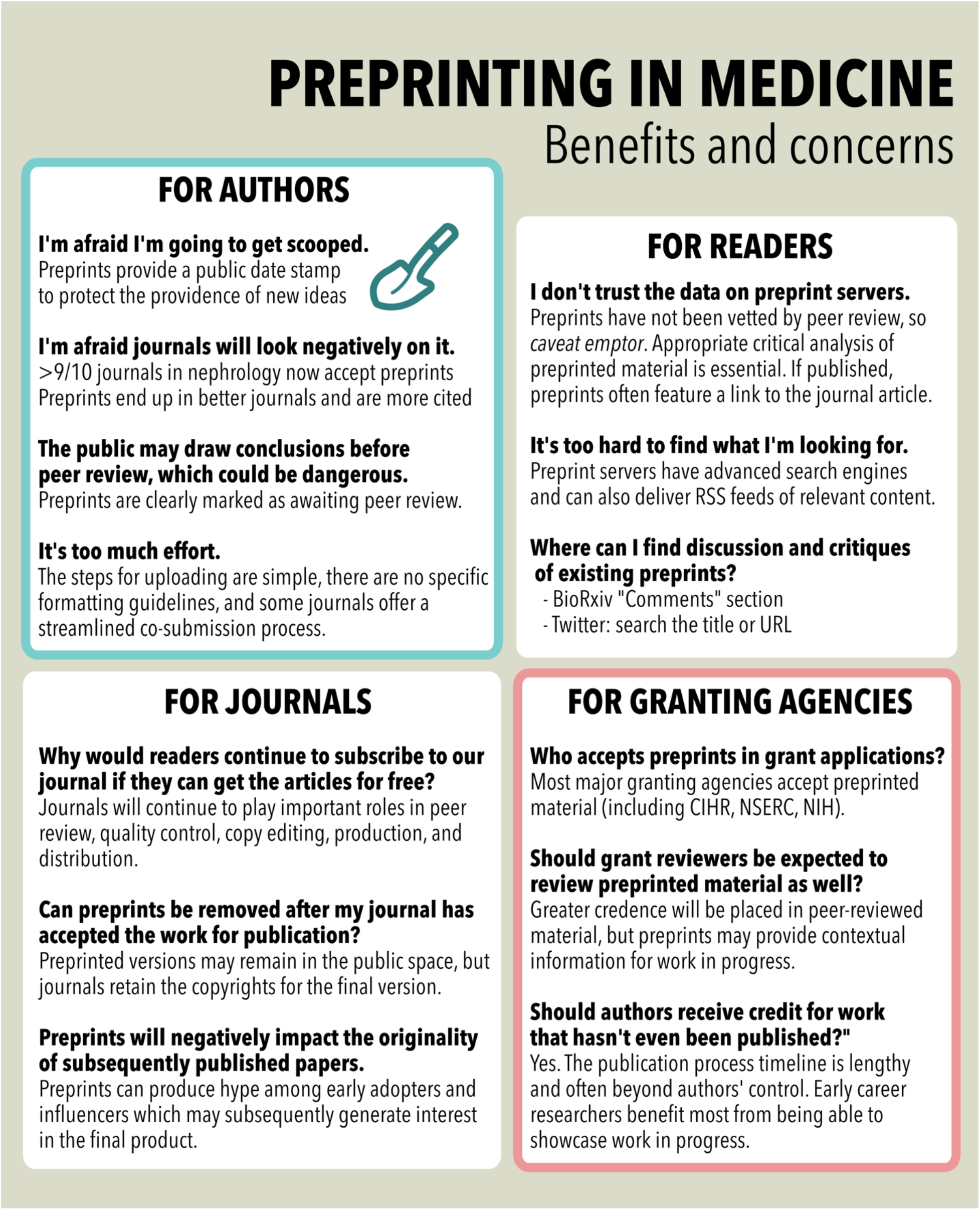
Summary of main points.

To better characterize how preprints can be incorporated into nephrology research we started with delineating which journals accept preprints. We contacted the twenty top--ranked journals in nephrology to inquire about their policies about accepting submissions that had been uploaded to preprint servers. Given their publication model of invited reviews, *Nature Reviews Nephrology* and *Advances in Chronic Kidney Disease* do not accept preprints (Table 1). Of the remaining 18 journals that publish primary research, 16 (89%) accept preprints. *American Journal of Kidney Disease* (AJKD) offered the following explanation for their decision to not accept preprints in email correspondence: “AJKD’s submission policies include the following requirement: Except by explicit, prior arrangement, manuscripts are considered for publication if the article or its key features (1) are not under consideration elsewhere, (2) have not already been disseminated in print or online, and (3) will not be disseminated in print or online prior to publication in AJKD. […] By this policy, a manuscript that had been uploaded as a preprint would not be eligible for consideration by AJKD (unless some prior arrangement had been made).” Similarly, the Journal of Renal Nutrition’s policy states that: “Except by explicit, prior arrangement, the *Journal of Renal Nutrition* will not consider for publication manuscripts that have been previously posted to preprint server, as this may jeopardize the journal’s double-- blind peer review practice.”

**Table 1.** Preprint policies reported by the top 20 journals in Nephrology.

| Journals that accept preprinted articles | Journals that do not accept preprinted articles |
| --- | --- |
| American Journal of Nephrology | Advances in Chronic Kidney Disease – *Invited reviews only* |
| BMC Nephrology | American Journal of Kidney Disease (AJKD) |
| Canadian Journal of Kidney Health and Disease (CJKHD) | Journal of Renal Nutrition |
| Clinical Journal of the American Society of Nephrology (CJASN) | Nature Reviews Nephrology – Invited reviews only |
| CKJ: Clinical Kidney Journal |  |
| Current Opinion in Nephrology and Hypertension – invited reviews only |  |
| International Journal of Nephrology and Renovascular Disease |  |
| Journal of the American Society of Nephrology (JASN) |  |
| Journal of Nephrology |  |
| Kidney International (KI) |  |
| Kidney International Reports (KIR) |  |
| Kidney International Supplements (KIS) |  |
| Nephron |  |
| Nephrology, Dialysis and Transplantation |  |
| Pediatric Nephrology |  |
| Seminars in Nephrology |  |

## Review & Discussion

### Preprint servers improve transparency and mitigate “scooping”

Preprints provide detailed accounts of work--in--progress and enable greater control over how research is disseminated. As all preprints have permanent, time--stamped digital object identifiers (DOIs), researchers can claim an indisputable stake and time stamp for discoveries. This DOI mitigates the risk of “scooping”– a seemingly intuitive concern held by those considering preprinting. However, in disciplines where preprinting is the standard, scooping is virtually absent as the providence of novel findings or concepts are part of the public domain^21^. In the traditional peer review model, established researchers serving as peer reviewers receive early access to yet unpublished results as part of the review process. Researchers with established expertise may receive less resistance from the peer review process for novel concepts. Preprint servers removes this competitive advantage by allowing all the opportunity to view results before formal publication.

### Preprint servers allow for early non--competitive peer review

Preprint journal clubs are becoming increasingly commonplace, serving as an exercise in critical analysis and exposure to the peer review process for journal club participants, while also providing valuable feedback to the authors to improve the paper before submission to the competitive publication process.^22,23^ The purpose of preprint servers is not to replace or undermine the importance of traditional peer review. Many publishers in the areas of biomedical and life sciences now encourage authors to upload their work to preprint servers via streamlined co--submission processes (i.e., via B2J and M2J). Eve Marder, deputy editor of eLife, described preprints as an opportunity to identify mistakes prior to final publication, expedite the availability of new results, and provide equitable access to the public^24^. Unfortunately, “We regret to inform you that your paper cannot be accepted for publication” is a phrase all researchers are familiar with. Authors are often not given an opportunity to respond or adapt their submission to the reviewer’s suggestions. Preprint servers may democratize access to early pre--submission peer review, as formerly, only those in institutions with internal reviews or mentors available for advice may have had access.

### Preprint servers allow for rapid access and dissemination

Some consider preprinting a compromise of rigor for greater speed of dissemination. However, the primary goal is not necessarily to disseminate, but rather to prompt in--depth discussion and feedback while formal peer review is ongoing. Rather than a limited number of reviewers critiquing a body of work, preprints facilitate ongoing evolution of a paper with widespread evaluation by the broader research community. Conference presentations have long served as the venue for delivery of unpublished results. In a similar fashion, conference proceedings have provided early access to results before peer reviewed publication, while providing authors with an opportunity for comments from potential peer reviewers. With social media, slides from such presentations are now often broadly distributed. Preprint servers can now serve as a formal, democratic, globally accessible venue for early access to results. In the context of outbreak science were rapid dissemination maybe of great importance, preprint servers may have specific benefits.^25^ During the outbreak of the 2019 coronavirus (COVID--19) pandemic over 500 preprints were uploaded to either bioRxiv or medRxiv between December 1, 2019 (date of the first reported case) and March 16, 2020.

### Researchers publishing preprints may identify new collaborations increasing the impact of their results

External validation greatly increases confidence in the validity of results. Two researchers publishing preprints addressing a similar question may realize that the impact of their results may be much greater as a single publication than separate reports.

### Preprint servers allow for citation of early work in grant funding applications

Major North American health research funding bodies (i.e., Canadian Institutes of Health Research, Natural Sciences and Engineering Research Council of Canada and the United States National Institutes of Health) have issued statements encourage preprinting in this context.

Preprinting may be particularly helpful for early career researchers, whose earlier research may have not had the time to percolate through the peer review process.^26^

### Preprint servers improve transparency

Preprinting is one modality that benefits from the collective criticism of the online scientific community to assist with peer review prior to certifying the data before publication. Some journals are piloting the publication of peer review reports to increase transparency of the peer review process. As of late 2019, *eLife* and *EMBO Press* journals are making their peer review reports via the bioRxiv Transparent Reviews in Preprints (TRiP) project^39^. Select *Nature* journals are also now beginning to publish peer review reports^40^. While anonymous comments can be made on preprint servers, comments or suggestions are available for all to read and posters often make their identity known.

### Preprinting confers greater visibility and recognition

Three independent analyses of the largest biological science repository, bioRxiv, revealed that articles with preprints receive higher Altmetric scores and more citations than articles without a preprint^27,28,29^. Approximately two--thirds of bioRxiv preprints are eventually published, after a median time interval of 5.5 months^4^. Interest in a preprint paper, signaled by high download count, may influence editorial outcomes: journals with higher impact factors tend to publish papers with high preprint download count^4^. Still in its infancy, similar analyses of medRxiv are expected in the coming years.

### Specific challenges in medical research for preprint servers

Public interest in early access to research developments is particularly acute in medicine, potentially distinguishing medical research from other fields where preprinting is a reflexive part of the publication process^13^. By definition preprints have not undergone peer review, and could lead to spreading of flawed conclusions. Medical preprints could pose greater risk for harm if relevant but unsubstantiated claims are adopted by physicians, patients, caregivers, or the media^30^. Whereas research scientists are trained to identify potential pitfalls in unpublished preprints, there is debate about whether this holds true for those not accustomed to reading primary literature^30,31^. When it comes to health decisions, an individual’s health could be negatively impacted by misinterpretation of an unreviewed report.^32^ MedRxiv implemented a checkpoint in an attempt to minimize the risk of harm: prior to upload contributors are asked whether the preprint could be harmful to patients, which “may include, but is not limited to, studies describing dual--use research and work that challenges or could compromise accepted public health measures and advice regarding infectious disease transmission, immunization, and therapy”^33^. To our knowledge, there are no specific examples of medical preprints that have actualized this type of public harm to date.

In contrast, identification of flawed research on a preprint server can lead to correction before publication, preventing publication and subsequent retraction. For example, a preprint titled “Uncanny similarity of unique inserts in the 2019--nCoV spike protein to HIV--1 gp120 and Gag”^34^ were criticized on Twitter and on bioRxiv, leading to its voluntary withdrawal by the authors less than 2 days after it was posted, before it gained any important media coverage^35^. In contrast, a correspondence describing asymptomatic coronavirus transmission published in the *New England Journal of Medicine* after ultra--rapid peer review was openly criticized in *Science* less than 4 days after initial publication^36^ when it was revealed that incomplete patient history had led to flawed conclusions. The article garnered widespread media coverage and informed the opinion of prominent public health officials^36^, influencing quarantines and travel bans^37^ prior to being corrected online. Another rapidly peer reviewed article describing the healthcare working conditions in Wuhan, China published in *Lancet Global Health* was swiftly retracted two days after publication due to false claims that it presented first--hand accounts^38^. Errors identified in the preprint stage can therefore help avoid the spread of misinformation (instead of a true retraction tracked by RetractionWatch, for example).

## Conclusion

The vast majority of nephrology journals currently accept primary research articles that have been submitted to publicly available preprint servers, and the number of nephrology preprints is increasing exponentially. There is a significant lag between study completion and publication, and results are often informally available at conference proceedings long before formal publication. Speed of research dissemination can be of great importance, especially in the context of outbreak science. The traditional model of peer review is evolving to promote transparency and equity of the scientific publication process. In medicine, safety concerns exist regarding making scientific results publically available prior to traditional peer review. Overall, with appropriate safeguards in place it appears that benefits of preprint servers outweigh the risks, and their use is likely to continue to grow.

## Data Availability

The data is included as Supplementary Information.

## Acknowledgements

The authors declare no conflicts of interest.

## Notes

### Competing Interest Statement

The authors have declared no competing interest.

### Funding Statement

No external funding was used to generate this report.

## References

1. Clarke, R. F. & Clarke, H. G. Recent Trends of Biomedical Offprint Distribution. Bull. Med. Libr. Assoc. 54, 38–41 (1966).

2. Cobb, M. The prehistory of biology preprints: A forgotten experiment from the 1960s. PLOS Biol. 15, e2003995 (2017).

3. Berg, J. M. et al. Preprints for the life sciences. Science 352, 899–901 (2016).

4. Abdill, R. J. & Blekhman, R. Tracking the popularity and outcomes of all bioRxiv preprints. eLife 8, (2019).

5. bioRxiv.org -- the preprint server for Biology. https://www.biorxiv.org/.

6. Sweet, D. Merge ahead: Introducing direct submission from bioRxiv. http://crosstalk.cell.com/blog/merge-ahead-direct-submission-from-biorxiv.

7. McConnell, J. & Horton, R. Having electronic preprints is logical. BMJ 316, 1907 (1998).

8. Relman, A. S. The Ingelfinger Rule. N. Engl. J. Med. 305, 824–826 (1981).

9. New CMAJ and CMAJ Open policy permitting preprints | CMAJ. https://www-cmaj-ca.proxy.bib.uottawa.ca/content/191/27/E752.

10. Rawlinson, C. & Bloom, T. New preprint server for medical research. BMJ 365, (2019).

11. Peiperl, L. & Editors, on behalf of the P. M. Preprints in medical research: Progress and principles. PLOS Med. 15, e1002563 (2018).

12. New Preprint Server for the Health Sciences Announced Today | BMJ. https://www.bmj.com/company/newsroom/new-preprint-server-for-the-health-sciences-announced-today/.

13. Maslove, D. M. Medical Preprints—A Debate Worth Having. JAMA 319, 443–444 (2018).

14. da Silva, J. A. T. The preprint debate: What are the issues? Med. J. Armed Forces India 74, 162–164 (2018).

15. Colbert, G. B. et al. The Social Media Revolution in Nephrology Education. Kidney Int. Rep. 3, 519–529 (2018).

16. Ting DK, Boreskie P, Luckett--Gatopoulos S, Gysel L, Lanktree MB, Chan TM (2020) Quality Assurance Techniques for Online Educational Resources in Medicine: A Rapid Review. Seminars in Nephrology. In press.

17. Covidence. https://community.cochrane.org/help/tools--and--software/covidence.

18. Topf, J. M. et al. Twitter--Based Journal Clubs: Additional Facts and Clarifications. J. Med. Internet Res. 17, (2015).

19. Renal Fellow Network -- For Fellows, By Fellows. Renal Fellow Network https://www.renalfellow.org/.

20. AJKD Blog. AJKD Blog https://ajkdblog.org/.

21. Bourne, P. E., Polka, J. K., Vale, R. D. & Kiley, R. Ten simple rules to consider regarding preprint submission. PLOS Comput. Biol. 13, e1005473 (2017).

22. Avasthi, P., Soragni, A. & Bembenek, J. N. Point of View: Journal clubs in the time of preprints. eLife https://elifesciences-org.proxy.bib.uottawa.ca/articles/38532. (2018) doi:10.7554/eLife.38532.

23. Casadevall, A. & Gow, N. Using Preprints for Journal Clubs. mBio 9, (2018).

24. Marder, E. Beyond scoops to best practices. eLife 6, e30076 (2017).

25. Preprints: An underutilized mechanism to accelerate outbreak science. https://journals.plos.org/plosmedicine/article?id=10.1371/journal.pmed.1002549.

26. Sarabipour, S. et al. On the value of preprints: An early career researcher perspective. PLOS Biol. 17, e3000151 (2019).

27. The effect of bioRxiv preprints on citations and altmetrics | bioRxiv. https://www.biorxiv.org/content/10.1101/673665v1.

28. Serghiou, S. & Ioannidis, J. P. A. Altmetric Scores, Citations, and Publication of Studies Posted as Preprints. JAMA 319, 402–404 (2018).

29. Fu, D. Y. & Hughey, J. J. Releasing a preprint is associated with more attention and citations for the peer-reviewed article. eLife 8, (2019).

30. Preprints could promote confusion and distortion. https://www.nature.com/articles/d41586-018-05789-4.

31. Sarabipour, S. Preprints are good for science and good for the public. Nature http://www.nature.com/articles/d41586-018-06054-4 (2018) doi:10.1038/d41586-018-06054-4.

32. Bauchner, H. The Rush to Publication: An Editorial and Scientific Mistake. JAMA 318, 1109–1110 (2017).

33. Advancing the sharing of research results for the life sciences. https://www.medrxiv.org/content/about-medrxiv.

34. Pradhan, P. et al. Uncanny similarity of unique inserts in the 2019-nCoV spike protein to HIV-1 gp120 and Gag. bioRxiv 2020.01.30.927871 (2020) doi: 10.1101/2020.01.30.927871.

35. Quick retraction of coronavirus paper was good moment for science. STAT https://www.statnews.com/2020/02/03/retraction-faulty-coronavirus-paper-good-moment-for-science/ (2020).

36. Kupferschmidt, K. 2020. Study claiming new coronavirus can be transmitted by people without symptoms was flawed. Science | AAAS https://www.sciencemag.org/news/2020/02/paper-non-symptomatic-patient-transmitting-coronavirus-wrong (2020).

37. Johnson, C. Y. & Lena H. S. Key evidence for coronavirus spread is flawed as public health decisions loom. Washington Post https://www.washingtonpost.com/health/2020/02/04/key-evidence-coronavirus-spread-is-retracted-public-health-decisions-loom/ (2020).

38. Oransky, A. I. Lancet journal retracts letter on coronavirus because authors say it “was not a first-hand account” after all. Retraction Watch https://retractionwatch.com/2020/02/27/lancet-journal-retracts-letter-on-coronavirus-because-authors-say-it-was-not-a-first-hand-account-after-all/ (2020).

39. Transparent review in preprints. Cold Spring Harbor Laboratory https://www.cshl.edu/transparent-review-in-preprints/ (2019).

40. Nature will publish peer review reports as a trial. Nature 578, 8–8 (2020).

